# AORTA Gene: Polygenic prediction improves detection of thoracic aortic aneurysm

**DOI:** 10.1101/2023.08.23.23294513

**Authors:** James P. Pirruccello, Shaan Khurshid, Honghuang Lin, Lu-Chen Weng, Siavash Zamirpour, Shinwan Kany, Avanthi Raghavan, Satoshi Koyama, Ramachandran S. Vasan, Emelia J. Benjamin, Mark E. Lindsay, Patrick T. Ellinor

**Affiliations:** Division of Cardiology, University of California San Francisco, San Francisco, California, USA; Institute for Human Genetics, University of California San Francisco, San Francisco, California, USA; Bakar Computational Health Sciences Institute, University of California San Francisco, San Francisco, California, USA; Cardiology Division, Massachusetts General Hospital, Boston, Massachusetts, USA; Cardiovascular Research Center, Massachusetts General Hospital, Boston, Massachusetts, USA; Cardiovascular Disease Initiative, Broad Institute of MIT and Harvard, Cambridge, Massachusetts, USA; Harvard Medical School, Boston, Massachusetts, USA; Framingham Heart Study, Boston University and National Heart, Lung, and Blood Institute, Framingham, Massachusetts, USA; Department of Medicine, University of Massachusetts Chan Medical School, Worcester, Massachusetts, USA; School of Medicine, University of California San Francisco, San Francisco, California, USA; Department of Cardiology, University Heart and Vascular Center Hamburg-Eppendorf, Hamburg, Germany; Laboratory for Cardiovascular Genomics and Informatics, RIKEN Center for Integrative Medical Sciences, Kanagawa, Japan; Department of Medicine, Cardiology and Preventive Medicine Sections, Boston Medical Center, Boston University Chobanian and Avedisian School of Medicine, Boston, Massachusetts, USA; Epidemiology Department, Boston University School of Public Health, Boston, Massachusetts, USA; Thoracic Aortic Center, Massachusetts General Hospital, Boston, Massachusetts, USA

**Author notes:** **Correspondence**: James P. Pirruccello, MD University of California San Francisco, Division of Cardiology, 555 Mission Bay Blvd South #3118, San Francisco, CA 94158; Patrick T. Ellinor, MD, PhD, Massachusetts General Hospital, Cardiology Division, 55 Fruit Street, Boston, MA 02114.

**Keywords:** Ascending aorta, polygenic score, UK Biobank, *All of Us*, Framingham Heart Study, Mass General Brigham

## Abstract

**Background:** Thoracic aortic disease is an important cause of morbidity and mortality in the US, and aortic diameter is a heritable contributor to risk. Could a polygenic prediction of ascending aortic diameter improve detection of aortic aneurysm?

**Methods:** Deep learning was used to measure ascending thoracic aortic diameter in 49,939 UK Biobank participants. A genome-wide association study (GWAS) was conducted in 39,524 participants and leveraged to build a 1.1 million-variant polygenic score with *PRScs-auto*. Aortic diameter prediction models were built with the polygenic score (“AORTA Gene”) and without it. The models were tested in a held-out set of 4,962 UK Biobank participants and externally validated in 5,469 participants from Mass General Brigham Biobank (MGB), 1,298 from the Framingham Heart Study (FHS), and 610 participants from *All of Us*.

**Results:** In each test set, the AORTA Gene model explained more of the variance in thoracic aortic diameter compared to clinical factors alone: 39.9% (95% CI 37.8-42.0%) vs 29.2% (95% CI 27.1-31.4%) in UK Biobank, 36.5% (95% CI 34.4-38.5%) vs 32.5% (95% CI 30.4-34.5%) in MGB, 41.8% (95% CI 37.7-45.9%) vs 33.0% (95% CI 28.9-37.2%) in FHS, and 34.9% (95% CI 28.8-41.0%) vs 28.9% (95% CI 22.9-35.0%) in *All of Us*. AORTA Gene had a greater AUROC for identifying diameter ≥4cm in each test set: 0.834 vs 0.765 (P=7.3E-10) in UK Biobank, 0.808 vs 0.767 in MGB (P=4.5E-12), 0.856 vs 0.818 in FHS (P=8.5E-05), and 0.827 vs 0.791 (P=7.8E-03) in *All of Us*.

**Conclusions:** Genetic information improved estimation of thoracic aortic diameter when added to clinical risk factors. Larger and more diverse cohorts will be needed to develop more powerful and equitable scores.

## Introduction

Thoracic aortic disease is an important cause of morbidity and mortality^1, 2^. Ascending thoracic aortic enlargement is a well-established risk factor for ascending aortic dissection^3, 4^. Indeed, the majority of dissections occur in individuals with aortic diameter ≥4cm, including more than 90% in the International Registry of Aortic Dissection and 100% in Kaiser^4, 5^. Contemporary guidelines do not describe a role for population screening of thoracic aortic aneurysm^6^, and universal imaging would likely be impractical.

These limitations have spurred the development of clinical risk scores to identify individuals who are likely to have aortic enlargement on confirmatory imaging^7–9^. These scores have been shown to explain approximately 30% of the variance in ascending aortic diameter, with an area under the receiver operator characteristic curve (AUROC) near 0.78 for detecting individuals with diameter ≥4cm. Population genetics studies suggest that ascending aortic diameter is highly heritable, with a proportion of variance attributable to common genetic factors of over 60%^10^. We therefore sought to assess whether incorporating polygenic risk might improve estimation of ascending aortic diameter over clinical factors alone.

## Methods

### Study design

Model development and internal validation were conducted in UK Biobank, with external validation within the *All of Us* cohort, the Mass General Brigham Biobank (MGB), and the Framingham Heart Study (FHS). This study was reported in accordance with the transparent reporting of a multivariable prediction model for individual prognosis or diagnosis (TRIPOD) statement^11^.

Study protocols complied with the tenets of the Declaration of Helsinki. All UK Biobank participants provided written informed consent^12^, and only those participants who had not withdrawn consent as of March 7, 2023 were analyzed. The UK Biobank analyses were considered exempt by the UCSF Institutional Review Board (IRB), #22-37715. UK Biobank analyses were conducted under application #41664. Each *All of Us* biobank participant provided written informed consent^13^. The *All of Us* analyses were considered exempt by the UCSF IRB, #22-37715. The MGB study protocols were approved with a waiver of informed consent by the Mass General Brigham IRB. All FHS participants provided written informed consent, and the FHS imaging analyses were previously approved by the IRB of the Boston University Medical Center^14^. The FHS analyses were also approved by the MGB IRB.

### Study populations

In UK Biobank participants, ascending aortic diameter was measured from magnetic resonance imaging (MRI) using a previously described deep learning model^15^. Model development and internal validation were conducted in UK Biobank. External validation of the models was pursued in MGB participants with ascending aortic diameter measured from TTE for clinical indications^16^; in FHS participants with ascending aortic diameter measured for research purposes from non-contrast computed tomography (CT)^14^; and in *All of Us* biobank participants who had clinical aortic diameter measurements from transthoracic echocardiography (TTE)^13^. In MGB, race or ethnicity was extracted from the electronic health record^16^ and the validation analyses were repeated in individuals identified as Black.

### Modality-specific differences

The clinical approach to measuring ascending aortic diameter differs by modality. UK Biobank MRI-based aortic diameter measurements were derived from the blood pool diameter only, incorporating neither aortic wall. For TTE, used in the MGB and *All of Us* cohorts, standard measurements use the “leading edge to leading edge” approach, incorporating one wall. For non-contrast CT measurements, used in FHS, both walls are incorporated into the diameter.

### Splitting the UK Biobank for training and validation

49,939 UK Biobank participants with MRI measurements of aortic diameter were randomly assigned a number from 0-999. Those from 0-799 contributed to the genome-wide association study (GWAS) used to produce the polygenic score (N=39,524). Those from 800-999 related within 3 degrees of kinship to the 0-799 group were excluded to avoid polygenic score overfitting (N=557). Those remaining from 0-899 contributed to the development of non-genetic models (N=44,420). Those from 800-899 were included in the derivation of weights for the polygenic score component of the AORTA Gene combined clinical and genetic model (N=4,896). Those ≥900 were analyzed for validation (N=4,962; **Figure 1**).

**Figure 1:**
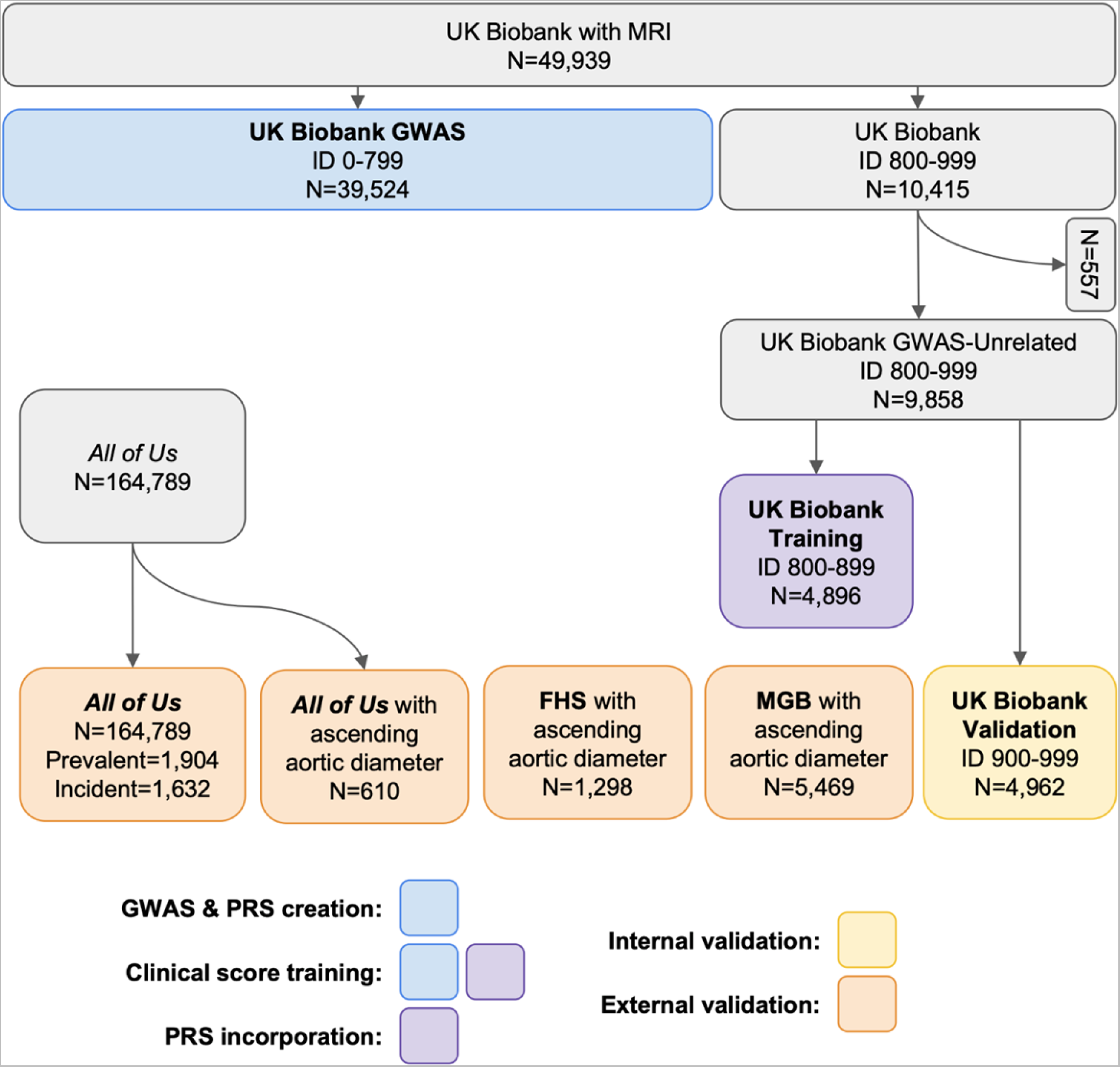
Study cohorts. The sample diagram depicts participant subsets. UK Biobank participants with MRI data were split by random ID into a GWAS group (blue, N=39,524) and a residual group. Individuals in the residual group related within 3 degrees of kinship were removed (N=557), and the residual group was then split into a training group (purple, N=4,896) and an internal validation group (yellow, N=4,962). As depicted in the legend, the GWAS and PRS development was conducted in the GWAS group (blue, N=39,524). Clinical model development was conducted in the GWAS and training groups (blue and purple, N=44,420). The PRS was incorporated into the clinical score using a linear model to produce the AORTA Gene model in the training group (purple, N=4,896). All models were validated in the internal validation set (yellow, N=4,962) and the external validation sets (orange): MGB, FHS, and *All of Us*. MGB: Mass General Brigham Biobank. FHS: Framingham Heart Study. MRI: magnetic resonance imaging. GWAS: genome-wide association study.

### Thoracic aortic aneurysm in *All of Us*

Within *All of Us*, thoracic aortic aneurysm was defined as the first occurrence of SNOMED codes 433068007, 74883004, or 426948001; ICD10CM codes of I71.01, I71.1, I71.2, or I77.810; ICD9CM codes of 441.01, 441.1, 441.2, or 447.71; ICD10PCS codes of 02RX0JZ or 02RW0JZ; the ICD9Proc code 39.73; or CPT4 codes 33863, 75957, 33880, 75956, or 33881. Prevalent analyses were conducted based on the presence of disease labels prior to the time of enrollment. Time-to-event analyses were conducted beginning at the time of enrollment and censored on July 1, 2022 using the R *survival* package, excluding participants whose diagnoses occurred prior to enrollment. Any tabular data with an *All of Us* sample count <20 was modified in accordance with *All of Us* policies.

### Genome-wide association study and polygenic scores

Common genetic contributions to ascending aortic diameter in the 39,524 GWAS participants were discovered using REGENIE v3.2.7^17^ with imputed variants provided by UK Biobank^18, 19^. GWAS covariates included age, age^2^, sex, the MRI serial number, the genotyping array, and the first ten principal components of ancestry. The summary statistics were trimmed to the ∼1.1 million UK Biobank-specific variants within HapMap3 identified by the *PRScs* authors, and underwent Bayesian weighting by *PRScs-auto* on its default settings to produce polygenic score weights^20^.

The polygenic score was applied to all UK Biobank participants with imputed genetic data^19^. Variants with imputation quality <0.3 were removed.

The polygenic score was applied to all MGB participants with imputed genetic data^16, 21^ with imputation into the TOPMed panel^22^.

The polygenic score was applied to FHS participants with CT and genetic data available. For FHS, genotyping was done on the Affymetrix GeneChip 500K Array Set & 50K Human Gene Focused Panel (Affymetrix, Santa Clara, CA, USA). Variants with call rate <97% or Hardy–Weinberg equilibrium P<1E-06 were excluded. The remaining variants were then imputed to the 1000 Genomes Project phase I release 3 panel by MACH version 1.0^23, 24^. Variants with imputation quality <0.3 were removed.

The polygenic score was applied to *All of Us* participants with whole genome sequencing data available in release “R7”^25^.

Within each set of participants, the polygenic score was residualized for the first 20 principal components of ancestry (except for FHS, where 10 were used due to availability). The polygenic score in each group was zero-centered and scaled by the standard deviation from the UK Biobank training set: for studies that applied the average polygenic score (UK Biobank, *All of Us*) the standard deviation was 3.4E-07, and for those that applied the summed polygenic score (MGB, FHS) the standard deviation was 0.38.

### Ascending aortic diameter model training

Hierarchical group least absolute shrinkage and selection operator models were built to estimate ascending aortic diameter using the R package *glinternet*^26^. Three models were constructed with *glinternet* v1.0.11 using the following independent variables: (a) age, age^2^, and sex; (b) age, age^2^, sex, and polygenic score; (c) age, age^2^, body mass index (BMI), heart rate, systolic and diastolic blood pressure, height, weight, sex, and a history of diabetes, hypertension, or hyperlipidemia. A fourth model (d) was constructed by taking a linear combination of the model (c) and the polygenic score. The weighting for model (c) was estimated in all participants randomly assigned to 0-899, while the weighting for model (b) and the linear combination for model (d) were estimated in those assigned to 800-899 to avoid overlap with the GWAS cohort. For ease of reference, we label model (c) “AORTA Score” (AORTA: optimized regression for thoracic aneurysm) and model (d) “AORTA Gene”.

### Statistical analysis

All statistical significance tests were two-sided, with significance defined as P < 0.05, and performed in R 4.2.3 unless otherwise stated. For visualization, scores were plotted against measured thoracic aortic diameter.

#### Outcomes

The primary outcome was correlation between the scores and ascending aortic diameter using linear models, expressed as R^2^ (variance explained). Secondary outcomes included tests of calibration, and the performance of the scores for identifying ascending aortic diameter ≥4.0cm; the clinical diagnosis of aneurysm was also tested in *All of Us*.

To assess calibration, each score was used as an independent variable in a linear model predicting aortic diameter that also had an intercept, permitting significance testing of both the slope and the intercept. Mean absolute error was also computed.

To assess the performance for identifying diameter ≥4cm, the AUROC was computed. Confusion matrices and their derived statistical measures were produced based on the presence or absence of aortic diameter ≥4cm and the presence or absence of predicted diameters above a fixed threshold determined in the training set for each score. The thresholds for the confusion matrix analysis were defined in the 4,896 participants in the training set.

AUROCs were compared using the DeLong test^27^. The continuous net reclassification index (NRI) was calculated following the formula of Pencina, *et al*^28^.

### Data and code availability

Computed scores are returned to UK Biobank, where data are made available by UK Biobank to researchers from research institutions with genuine research inquiries, following IRB and UK Biobank approval. FHS data are made available to researchers with approved research applications. The dbGAP study accession number used for FHS validation was #phv00076329.v1.p5 for ascending aortic diameter. MGB data are available to MGB investigators; external collaboration requests can be initiated through https://biobank.massgeneralbrigham.org/for-researchers. The complete set of AORTA Score covariates and their weights are available as R programs at github.com/carbocation/genomisc and the polygenic score weights will be available on the Polygenic Score Catalog (pgscatalog.org).

## Results

### Clinical characteristics

Among the 4,896 UK Biobank participants whose data contributed to training the genetic models, 2,489 were women and 2,407 were men. The ascending aortic diameter was 3.05 ± 0.31cm for women and 3.34 ± 0.34cm for men. The 4,962 internal validation set participants had similar measurements (**Table 1**; **Figure 1**). The MGB validation set consisted of 5,469 participants with genetic data and aortic diameter measured from TTE. The FHS validation set consisted of 1,298 participants with genetic data and aortic diameter measured from non-contrast CT. The *All of Us* validation set consisted of 610 participants 40 years and older with genetic data and aortic diameter measured from TTE; 60% identified as female and 88% as having non-Hispanic white ethnicity.

**Table 1:** Participant characteristics. All values are mean (standard deviation) or count (percent of total). BPM: beats per minute. MGB: Mass General Brigham. BMI: body mass index. MRI: magnetic resonance imaging. CT: computed tomography. TTE: transthoracic echocardiography.

|  | UK Biobank<br>MRI Training | UK Biobank<br>MRI Internal<br>Validation | MGB TTE<br>External<br>Validation | FHS CT<br>External<br>Validation | *All of Us* TTE<br>External<br>Validation | *All of Us*<br>Aneurysm<br>External<br>Validation |
| --- | --- | --- | --- | --- | --- | --- |
| **N** | 44,420 | 4,962 | 5,469 | 1,298 | 610 | 164,789 |
| **Men** | 21,053 (47.4%) | 2,344 (47.2%) | 2,580 (47.2%) | 617 (47.5%) | 246 (40.3%) | 70,861 (43.0%) |
| **Women** | 23,367 (52.6%) | 2,618 (52.8%) | 2,889 (52.8%) | 681 (52.5%) | 364 (59.7%) | 93,928 (57.0%) |
| **Age (years)** | 65.2 (7.7) | 65.4 (7.7) | 61.1 (14.4) | 59.8 (9.0) | 66.1 (11.0) | 60.3 (10.9) |
| **Medical history** |  |  |  |  |  |  |
| Hypertension | 13,299 (29.9%) | 1,501 (30.2%) | 4,535 (82.9%) | 532 (41.0%) | 426 (69.8%) | 57,845 (35.1%) |
| Hyperlipidemia | 9,644 (21.7%) | 1,103 (22.2%) | 942 (17.2%) | 257 (19.8%) | 155 (25.4%) | 6,379 (3.9%) |
| Diabetes | 1,277 (2.9%) | 160 (3.2%) | 1,750 (32.0%) | 115 (8.9%) | 210 (34.4%) | 25,568 (15.5%) |
| **Heart rate (bpm)** | 68 (11) | 68 (11) | 76 (15) | 65 (11) | 69 (11) | 72 (12) |
| **Systolic blood**<br>**pressure**<br>**(mmHg)** | 140 (18) | 141 (18) | 128 (17) | 126 (18) | 129 (18) | 129 (18) |
| **Diastolic blood**<br>**pressure**<br>**(mmHg)** | 79 (10) | 79 (10) | 75 (10) | 75 (9) | 77 (11) | 78 (11) |
| **Height (cm)** | 169 (9) | 169 (9) | 169 (10) | 168 (9) | 166 (10) | 168 (10) |
| **Weight (kg)** | 75.2 (14.7) | 75.4 (14.9) | 84.4 (21.2) | 80 (17.3) | 88.9 (24.9) | 84.5 (21.8) |
| **BMI (kg/m<sup>2</sup>)** | 26.3 (4.2) | 26.3 (4.3) | 29.5 (6.6) | 28.3 (5.2) | 32.0 (8.4) | 30.0 (7.3) |
| **Ascending**<br>**aortic diameter**<br>**(cm)** | 3.18 (0.35) | 3.19 (0.35) | 3.20 (0.48) | 3.45 (0.36) | 3.34 (0.45) | NA |
| **Aortic diameter**<br>**≥4cm** | 1,040 (2.3%) | 113 (2.3%) | 335 (6.1%) | 116 (8.9%) | 65 (10.7%) | NA |
| **Prevalent**<br>**aneurysm** | NA | NA | NA | NA | NA | 1,904 (1.2%) |
| **Incident**<br>**aneurysm** | NA | NA | NA | NA | NA | 1,632 (1.0%) |

### Model validation for aortic diameter

Testing calibration, the slope of the AORTA Gene model was statistically consistent with one in UK Biobank (P=0.2), MGB (P=1.0), FHS (P=0.6), and *All of Us* (P=0.4) (**Figure 2**). The intercept was consistent with zero in UK Biobank (P=0.2), MGB (P=0.4), and *All of Us* (P=0.8), but not FHS (intercept estimate 0.26cm, P=0.01). The same pattern was observed for the clinical AORTA Score. In contrast, the model incorporating only age, sex, and the polygenic score had a calibration slope inconsistent with one in most cohorts outside of UK Biobank (**eTable AA**).

**Figure 2:**
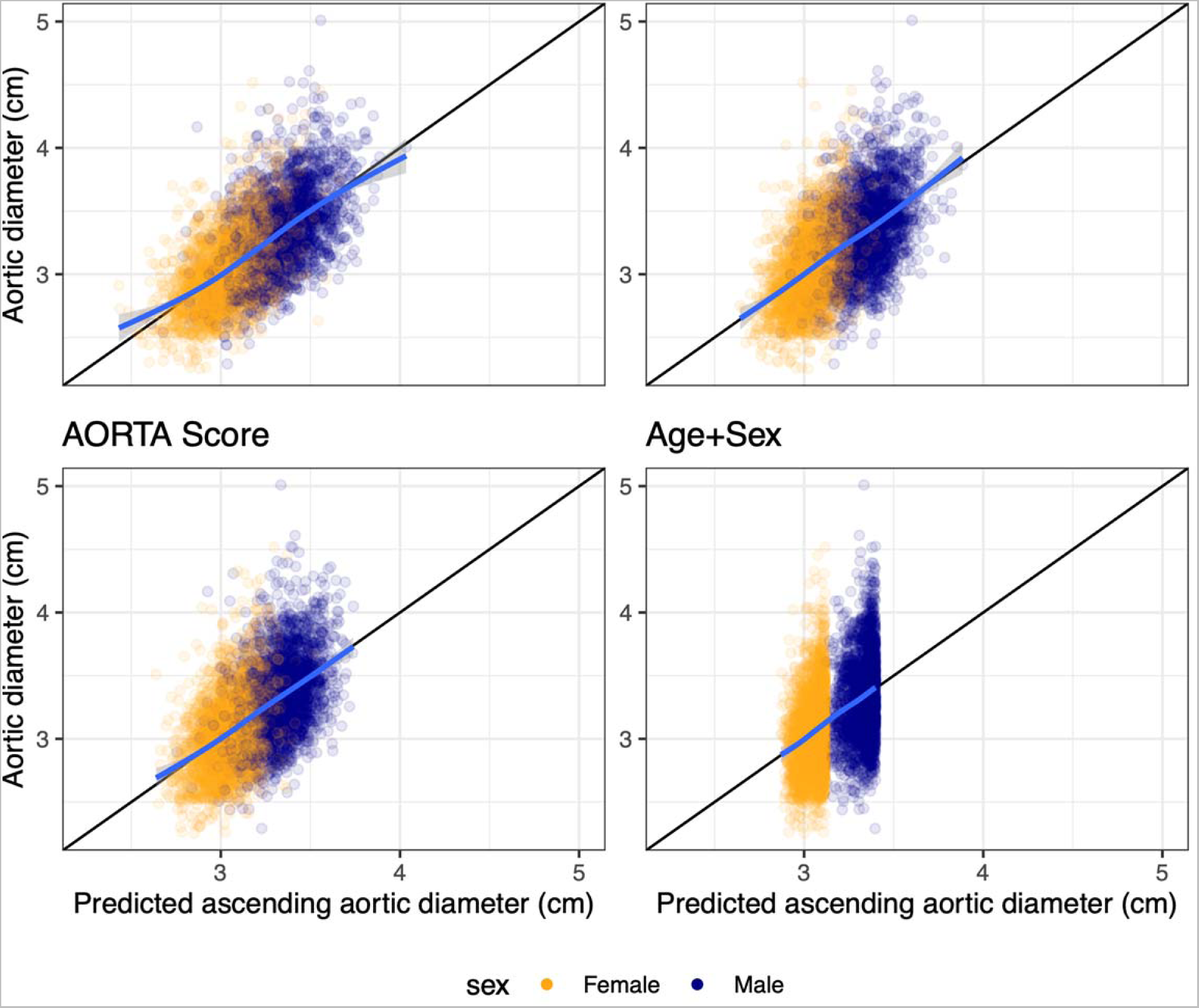
Score calibration curves in the UK Biobank validation set. Counterclockwise from top left: AORTA Gene, followed by the clinical AORTA Score, the age and sex model, and the age/sex/genetics model. The x axis represents the predicted ascending aortic diameter; the y axis represents the measured ascending aortic diameter; both are truncated at the same points so that the x and y axes have the same span. Each point represents one of the 4,962 UK Biobank internal validation set participants; orange points represent women while navy points represent men. The blue line shows the smoothed average value, and only extends along the x axis to the limits of the observed data. The black line shows the line of ideal calibration, where a 1 cm greater score would be met by a 1 cm greater aortic diameter. For all plotted scores, the slopes were statistically indistinguishable from one and the intercepts were statistically indistinguishable from zero as detailed in the Results.

The AORTA Gene model’s mean absolute error (MAE) for ascending aortic diameter was 0.212cm (95% CI 0.207-0.217cm) in UK Biobank, 0.282cm (95% CI 0.275- 0.289cm) in MGB, 0.344cm (95% CI 0.331-0.358cm) in FHS, and 0.293cm (95% CI 0.273-0.313cm) in *All of Us*. The clinical AORTA Score had an inferior MAE in all cohorts: 0.232cm (95% CI 0.227-0.237cm) in UK Biobank, 0.291cm (95% CI 0.284- 0.298cm) in MGB, 0.351cm (95% CI 0.337-0.365cm) in FHS, and 0.306cm (95% CI 0.285-0.327cm) in *All of Us*. The age/sex/genetics model had an MAE of 0.226cm (95% CI 0.221-0.231cm) in UK Biobank, 0.293cm (95% CI 0.286-0.300cm) in MGB, 0.346cm (95% CI 0.332-0.360cm) in FHS, and 0.318cm (95% CI 0.296-0.340) in *All of Us*.

The AORTA Gene model explained 39.9% of the variance in ascending aortic diameter (99% CI 37.8-42.0%; P=2.6E-551) in the UK Biobank validation set, 36.5% (95% CI 34.4-38.5%, P=2.4E-541) in MGB, 41.8% (95% CI 37.7-45.9%; P=3.5E-154) in FHS, and 34.9% (95% CI 28.8-41.0%; P=1E-58) in *All of Us* (**Figure 3**). In comparison, the clinical AORTA Score explained 29.3% of the variance in UK Biobank (99% CI 27.1- 31.4%), 32.5% in MGB (95% CI 30.4-34.5%), 33.0% (95% CI 28.9-37.2%) in FHS, and 28.9% (95% CI 22.9-35.0%) in *All of Us*. The age/sex/genetics model explained 31.8% (95% CI 29.6-33.9%) of variance in UK Biobank, 33.2% (95% CI 31.2-35.3%) in MGB, 33.3% (95% CI 29.1-37.5%) in FHS, and 31.0% (95% CI 24.9-37.1%) in *All of Us*.

**Figure 3:**
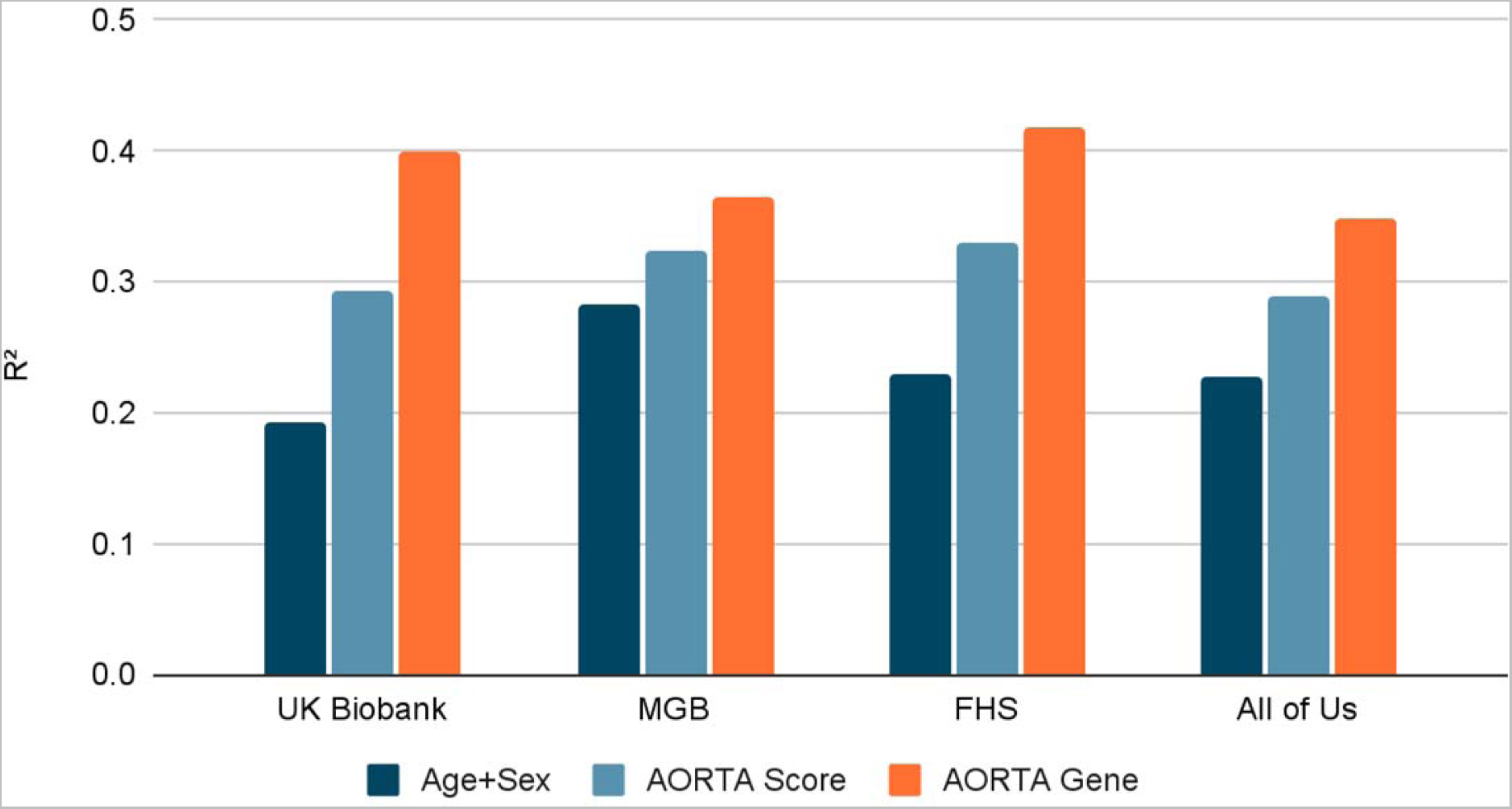
Variance in aortic diameter explained by the models. For each cohort, the variance in ascending aortic diameter explained by the model derived from age and sex (navy blue), the clinical model (AORTA Score, slate gray), and the clinical model incorporating the polygenic score (AORTA Gene, orange) is depicted for the internal validation set and the three external validation sets. MGB: Mass General Brigham Biobank. FHS: Framingham Heart Study.

### Model validation for a 4cm aortic diameter threshold

113 of the 4,962 internal validation set participants in UK Biobank had aortic diameter ≥4cm (2.3%), as did 335 of 5,469 MGB participants (6.1%), 116 of 1,298 FHS participants (8.9%), and 65 of 610 *All of Us* participants (10.7%).

The AUROC for detecting diameter ≥4cm for the AORTA Gene model was 0.834 (95% CI 0.798-0.869) in UK Biobank, 0.808 (95% CI 0.783-0.832) in MGB, 0.856 (95% CI 0.821-0.891) in FHS, and 0.827 (95% CI 0.776-0.878) in *All of Us* (**Figure 4**; **eTable BB**).

**Figure 4:**
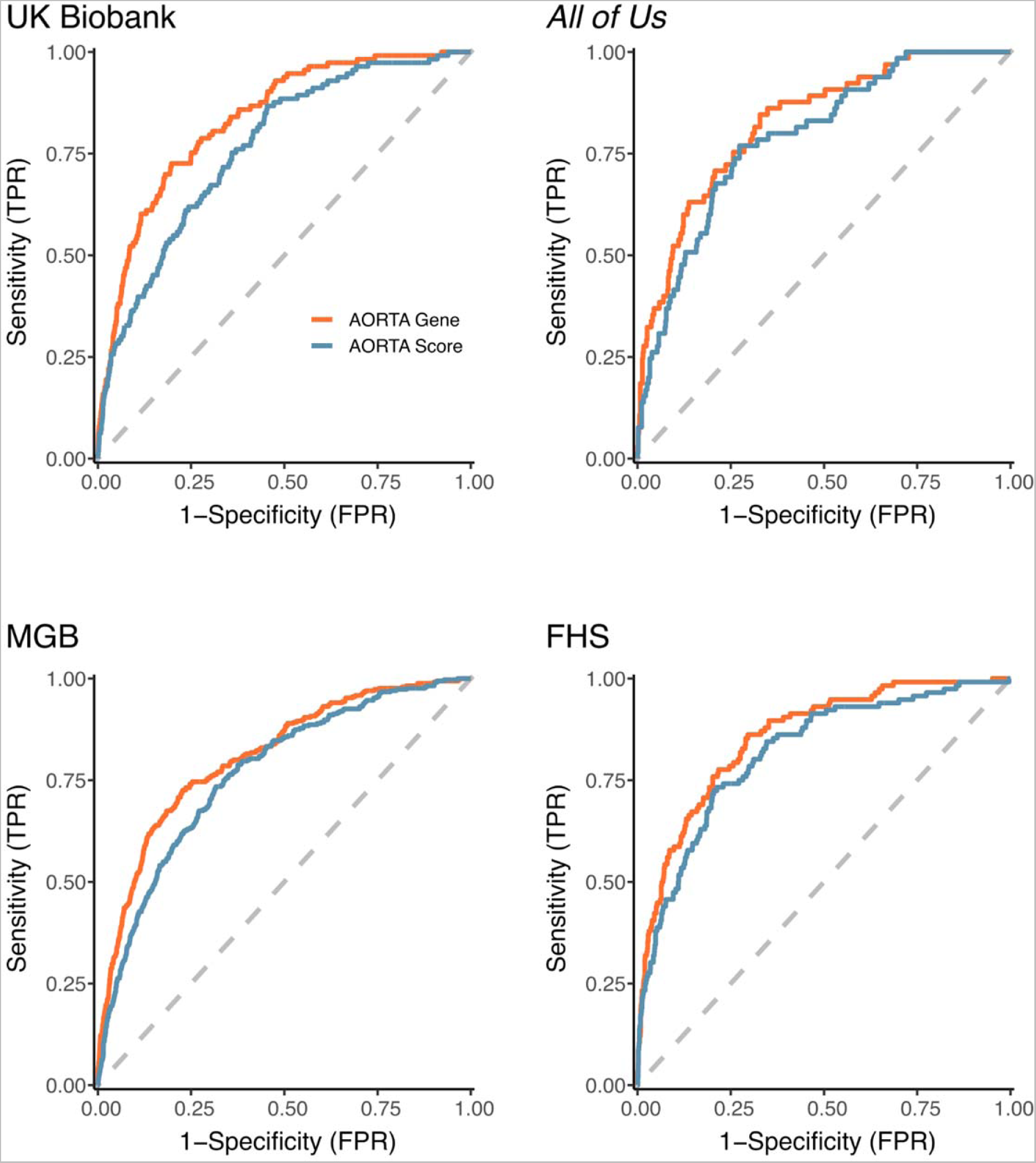
Receiver operator characteristic curves. Receiver operator characteristic (ROC) curves for the AORTA Gene model (red) and the clinical AORTA Score model (blue) in the validation cohorts. The dashed diagonal line represents the no-information baseline. Counterclockwise from top left: UK Biobank internal validation, MGB, FHS, and *All of Us*. The AUROC for detecting aortic diameter ≥4cm for the AORTA Gene model in these cohorts was, respectively, 0.834, 0.808, 0.856, and 0.827. The respective AUROC for the clinical AORTA Score was 0.765, 0.767, 0.818, and 0.791. MGB: Mass General Brigham Biobank. FHS: Framingham Heart Study.

For the clinical AORTA Score, the AUROC was 0.765 (95% CI 0.723-0.807) in UK Biobank, 0.767 (95% CI 0.741-0.792) in MGB, 0.818 (95% CI 0.776-0.859) in FHS, and 0.791 (95% CI 0.735-0.846) for *All of Us*. The AUROC of AORTA Gene was significantly greater in all cohorts: P=7.3E-10 against a null hypothesis of no difference in UK Biobank, P=4.5E-12 in MGB, P=8.5E-05 in FHS, and P=7.8E-03 in *All of Us* (**eTable CC**).

The age/sex/genetics model had an AUROC of 0.805 (95% CI 0.767-0.843) in UK Biobank, 0.805 (95% CI 0.781-0.829) in MGB, 0.819 (95% CI 0.781-0.856) in FHS, and 0.813 (95% CI 0.757-0.870) in *All of Us*. The AORTA Gene AUROC was significantly greater in UK Biobank (P=0.03) and FHS (P=3.9E-03), but not in MGB (P=0.7) or *All of Us* (P=0.5) (**eTable CC**).

The continuous NRI was positive for AORTA Gene over the clinical AORTA Score and the age/sex/genetics model in all cohorts (**eTable CC**). Decomposed into its NRI(event) and NRI(nonevent) components, AORTA Gene had a greater NRI(event) in all cohorts compared to the AORTA Score and the age/sex/genetics model. However, its NRI(nonevent) value was lower than that of the AORTA Score in MGB and UK Biobank, and lower than that of the age/sex/genetics model in MGB and *All of Us*.

### Score threshold demonstrations

To evaluate the consequences of various score thresholds whereby a portion of the population could be brought forward for confirmatory thoracic imaging, two thresholds were defined within the UK Biobank training set and applied in the validation sets: one targeted at the top 10% of the population, and the other designed to have a sensitivity of 90% for aortic diameter ≥4cm (**eTable BB**).

For the target top 10% AORTA Gene cutoff, 485 (9.8%) of 4,962 participants were above this threshold in UK Biobank, 640 (11.7%) of 5,469 in MGB, 99 (7.6%) of 1,295 in FHS, and 97 (15.9%) of 610 in *All of Us*. 59 of 113 UK Biobank participants with diameter ≥4cm were correctly classified as enlarged (sensitivity 52.2%), as were 162 of 335 in MGB (48.4%), 47 of 116 in FHS (40.5%), and 35 of 65 in *All of Us* (53.8%). The precision was 0.122 in UK Biobank, 0.253 in MGB, 0.475 in FHS, and 0.361 in *All of Us*, yielding respective F1 scores of 0.197, 0.332, 0.437, and 0.432 (**eTable BB**). A comparable threshold for the clinical AORTA Score flagged 484 (9.8%) of 4,962 UK Biobank participants, 722 (13.2%) of 5,469 in MGB, 92 (7.1%) of 1,295 in FHS, and 110 (18.0%) of 610 in *All of Us*. 41 of 113 UK Biobank participants with diameter ≥4cm were correctly classified as enlarged (sensitivity 36.3%) as were 143 of 335 in MGB (42.7%), 38 of 116 in FHS (32.8%), and 33 of 65 in *All of Us* (50.8%). The precision was 0.85 in UK Biobank, 0.198 in MGB, 0.413 in FHS, and 0.300 in *All of Us*, yielding respective F1 scores of 0.137, 0.271, 0.365, and 0.377 (**eTable BB**).

For the target 90% sensitive AORTA Gene cutoff, 1,737 (35.0%) of 4,962 UK Biobank participants were above this threshold, 1,943 (35.5%) of 5,469 in MGB, 336 (25.9%) of 1,295 in FHS, and 247 (40.5%) of 610 in *All of Us*. 92 of 113 UK Biobank participants with diameter ≥4cm were correctly classified as enlarged (sensitivity 81.4%), as were 258 of 335 in MGB (77.0%), 88 of 116 in FHS (75.9%), and 56 of 65 in *All of Us* (86.2%). The precision was 0.053 in UK Biobank, 0.133 in MGB, 0.262 in FHS, and 0.227 in *All of Us*, yielding respective F1 scores of 0.099, 0.227, 0.389, and 0.359 (**eTable DD**). A comparable threshold for the clinical AORTA Score flagged 2,006 (40.4%) of 4,962 UK Biobank participants, 2,194 (40.1%) of 5,469 in MGB, 405 (31.3%) of 1,295 in FHS, and 266 (43.6%) of 610 in *All of Us*. 87 of 113 UK Biobank participants with diameter ≥4cm were correctly classified as enlarged (sensitivity 77.0%), as were 264 of 335 in MGB (78.8%), 86 of 116 in FHS (74.1%), and 52 of 65 in *All of Us* (80.0%). The precision was 0.043 in UK Biobank, 0.120 in MGB, 0.212 in FHS, and 0.195 in *All of Us*, yielding respective F1 scores of 0.082, 0.209, 0.330, and 0.314 (**eTable DD**).

### Thoracic aortic aneurysm diagnosis in *All of Us*

In the 164,789 *All of Us* participants aged 40 or older with genetic data, 1,904 had an electronic health record (EHR)-based diagnosis of thoracic aortic aneurysm prior to enrollment. AORTA Gene’s AUROC was 0.760 (95% CI 0.750-0.771) compared to 0.739 (95% CI 0.728-0.750) for the AORTA Score (**eTable EE**), P=2.4E-16 against a null hypothesis of no difference (**eTable FF**). For incident disease (1,632 cases diagnosed after enrollment), AORTA Gene’s AUROC was 0.748 (95% CI 0.737-0.759) compared to 0.729 (95% CI 0.718-0.741) for the AORTA Score, P=9.5E-10 against a null hypothesis of no difference (**eTables EE-FF**).

### Analysis in Black individuals in MGB

In MGB, 340 individuals were identified as Black through the electronic health record^16^. In these individuals, AORTA Gene had an intercept consistent with zero (P=0.5) and a slope consistent with one (P=0.5). The MAE was 0.297cm (95% CI 0.272-0.321cm), and the model explained 34.8% of variance in ascending aortic diameter (95% CI 26.7- 43.0%). In comparison, for the clinical AORTA Score these values were 0.294cm (95% CI 0.269-0.319cm) and 34.3% (95% CI 26.2-42.5%) (**eTable AA**). The AUROC for detecting diameter ≥4cm for the AORTA Gene model was 0.858 (**eTable BB**), nominally greater than that of the clinical AORTA Score (AUROC 0.820, P=0.04 for the difference, **eTable CC**), with a positive continuous NRI (0.473).

## Discussion

Ascending thoracic aortic diameter has a SNP heritability estimated to be greater than 60%^10^, which makes the incorporation of common-variant genetic data for the presymptomatic detection of individuals with aortic enlargement or aneurysm conceptually appealing. But whether aortic polygenic scores encode information not already latently captured through correlation with clinical covariates was unknown. Here, we observed that AORTA Gene, which incorporated both an aortic polygenic score and clinical covariates, improved estimation of ascending aortic diameter and identification of individuals with diameter ≥4cm in four cohorts—UK Biobank, MGB, FHS, and *All of Us*—when compared to a previously validated model for aortic diameter built from the same clinical covariates.

For the estimation of aortic diameter as a quantitative measure, a clinical model built from the same covariates as the AORTA Score^9^ explained 29.3% of the variance in ascending aortic diameter in UK Biobank, 28.9% in *All of Us*, 32.5% in MGB, and 33.0% in FHS. The AORTA Gene model that additionally incorporated the aortic diameter polygenic score increased those values to 39.9% in UK Biobank, 34.9% in *All of Us*, 36.5% in MGB, and 41.8% in FHS. These represent a 10.6% absolute improvement in explained variance (36% relative improvement) in UK Biobank, 4% (12.3% relative improvement) in MGB, 8.8% (27% relative improvement) in FHS, and 6% (21% relative improvement) in *All of Us*. These substantive improvements from the polygenic score over a clinical model are comparable in magnitude to the addition of key clinical predictors over age and sex alone^9^.

Incorporation of the polygenic score also improved identification of individuals with clinically relevant aortic enlargement (i.e., diameter ≥4cm). The clinical model had an AUROC of 0.765 in UK Biobank, 0.791 in *All of Us*, 0.767 in MGB, and 0.818 in FHS; these respectively improved to 0.834, 0.827, 0.808, and 0.856 for AORTA Gene. The AUROC is independent of the choice of score threshold, so we also demonstrated two example thresholds: one based on the goal of being 90% sensitive for diameter ≥4cm, and another based on a scenario where there are resources to image up to 10% of the population. In both scenarios, for all four cohorts, sensitivity and specificity were modestly improved with the incorporation of the polygenic score. Interestingly, we also observed that, compared to the comprehensive clinical score, a simpler model consisting only of age, sex, and the polygenic score performed similarly. Because such a model can be estimated for any future time from birth, this may have implications for attempts at early risk stratification before the onset of clinical risk factors. We also noted that, for the task of identifying *All of Us* participants with an EHR-based diagnosis of aneurysm, all models had attenuated performance—likely in part due to limited ascertainment of thoracic aortic disease in current clinical practice. Nevertheless, even for this task, the AORTA Gene model remained superior to the clinical score (P=9.5E- 10).

We also note two limitations of the current model that point to future directions: first, despite a heritability of nearly 60%, the current polygenic score for thoracic aortic diameter derived from a GWAS of 39,524 participants added approximately 10% to the variance already explained by a clinical model, suggesting that it accounted for approximately 17% of the heritability of aortic diameter. In contrast, polygenic scores for height—built from GWAS of millions of participants—are nearly saturated^29^. Larger GWAS sample sizes are required to increase the variance explained by polygenic estimates of ascending aortic diameter. Second, polygenic scores derived in largely European-ancestry populations do not capture the full range of human genetic diversity, limiting their utility in individuals with more diverse genetic identities^30^. To examine this, we evaluated 340 individuals in MGB identified as Black in the electronic health record^16^. In these individuals, the addition of the polygenic score did not improve the continuous estimation of aortic diameter compared to the clinical score (R^2^ 34.8% for AORTA Gene vs 34.3% for the clinical AORTA Score), although it did nominally improve identification of individuals with diameter ≥4cm (AUROC 0.858 vs 0.820, P=0.04). We expect a key problem from incorporating genetic data to be one of inequity due to differential accrual of benefits to individuals with European genetic identities. Larger and more diverse GWAS are required to improve genetic predictions for people across diverse genetic ancestries.

Anticipation of the relevant clinical scenario for the application of polygenic scores is important for placing the current findings into context. The cost of acquiring a targeted set of images of the ascending thoracic aorta may be comparable that of obtaining genetic information. Therefore, we expect that the value of polygenic scores for primary prevention will be realized within the context of a healthcare system that incorporates genetic information as part of the prior probability across many diseases and risk factors, where the cost of one-time genotyping would be amortized over all downstream uses. In such a system, might there be a role, in principle, for the incorporation of a genetic estimator of thoracic aortic diameter into a comprehensive model to identify people who are likely to have ascending aortic enlargement? The results from the present analysis suggest so.

### Limitations

While all participants were incorporated into analyses without exclusion for race, ethnicity, or ancestry, the participants in UK Biobank, MGB, FHS, and *All of Us* with thoracic imaging predominantly had genetic identities similar to that of Europeans. None of the models described in this manuscript are anticipated to be effective for identifying individuals with aortic enlargement driven by rare pathogenic variants, which are not captured in polygenic scores constructed from common variants. The models were derived and tested in individuals over the age of 40 years. The attenuation in variance explained in the external cohorts compared to UK Biobank may be, in part, a consequence of cryptic relatedness causing overfitting to the UK Biobank cohort; reducing this discrepancy is an area of interest for future efforts. Future efforts are also needed to understand whether aortic prediction models can improve outcomes.

## Conclusion

Integrating genetic information into a validated clinical model improved estimation of ascending aortic diameter and the identification of individuals with thoracic aortic aneurysm.

## Data Availability

Computed scores are returned to UK Biobank, where data are made available by UK Biobank to researchers from research institutions with genuine research inquiries, following IRB and UK Biobank approval. FHS data are made available to researchers with approved research applications. The dbGAP study accession number used for FHS validation was #phv00076329.v1.p5 for ascending aortic diameter. MGB data are available to MGB investigators; external collaboration requests can be initiated through https://biobank.massgeneralbrigham.org/for-researchers. Upon publication, the complete set of AORTA Score covariates and their weights will be available as R programs at github.com/carbocation/genomisc and the polygenic score weights will be available on the Polygenic Score Catalog (pgscatalog.org).

## Acknowledgments

JPP had full access to all the data in the study and takes responsibility for the integrity of the data and the accuracy of the data analysis. JPP and PTE conceived of the study. JPP conducted the genetic analyses, developed the models, validated the models, and conducted bioinformatic analyses in the UK Biobank, MGB, and *All of Us*. HL validated the models in FHS. LCW applied the polygenic score in MGB. S Koyama performed PCA in MGB. JPP and PTE wrote the paper. All other authors contributed to the analysis plan or provided critical revisions.

## References

1. Howard DPJ, Banerjee A, Fairhead JF, Perkins J, Silver LE, Rothwell PM, Oxford Vascular Study. Population-based study of incidence and outcome of acute aortic dissection and premorbid risk factor control: 10-year results from the Oxford Vascular Study. Circulation. 2013;127:2031–2037.

2. Yamaguchi T, Nakai M, Yano T, Matsuyama M, Yoshino H, Miyamoto Y, Sumita Y, Matsuda H, Inoue Y, Okita Y, Minatoya K, Ueda Y, Ogino H. Population-based incidence and outcomes of acute aortic dissection in Japan. European Heart Journal Acute Cardiovascular Care. 2021;10:701–709.

3. Kim JB, Spotnitz M, Lindsay ME, MacGillivray TE, Isselbacher EM, Sundt TM. Risk of Aortic Dissection in the Moderately Dilated Ascending Aorta. J Am Coll Cardiol. 2016;68:1209–1219.

4. Solomon MD, Leong T, Sung SH, Lee C, Allen JG, Huh J, LaPunzina P, Lee H, Mason D, Melikian V, Pellegrini D, Scoville D, Sheikh AY, Mendoza D, Naderi S, Sheridan A, Hu X, Cirimele W, Gisslow A, Leung S, Padilla K, Bloom M, Chung J, Topic A, Vafaei P, Chang R, Miller DC, Liang DH, Go AS, Kaiser Permanente Northern California Center for Thoracic Aortic Disease. Association of Thoracic Aortic Aneurysm Size With Long-term Patient Outcomes: The KP-TAA Study. JAMA Cardiology [Internet]. 2022 [cited 2022 Oct 11];Available from: 10.1001/jamacardio.2022.3305

5. Pape LA, Tsai TT, Isselbacher EM, Oh JK, O’gara PT, Evangelista A, Fattori R, Meinhardt G, Trimarchi S, Bossone E, Suzuki T, Cooper JV, Froehlich JB, Nienaber CA, Eagle KA, International Registry of Acute Aortic Dissection (IRAD) Investigators. Aortic diameter >or = 5.5 cm is not a good predictor of type A aortic dissection: observations from the International Registry of Acute Aortic Dissection (IRAD). Circulation. 2007;116:1120–1127.

6. Isselbacher EM, Preventza O, Hamilton Black J, Augoustides JG, Beck AW, Bolen MA, Braverman AC, Bray BE, Brown-Zimmerman MM, Chen EP, Collins TJ, DeAnda A, Fanola CL, Girardi LN, Hicks CW, Hui DS, Schuyler Jones W, Kalahasti V, Kim KM, Milewicz DM, Oderich GS, Ogbechie L, Promes SB, Gyang Ross E, Schermerhorn ML, Singleton Times S, Tseng EE, Wang GJ, Woo YJ. 2022 ACC/AHA Guideline for the Diagnosis and Management of Aortic Disease: A Report of the American Heart Association/American College of Cardiology Joint Committee on Clinical Practice Guidelines. Circulation. 2022;146:e334–e482.

7. Mori M, Gan G, Deng Y, Yousef S, Weininger G, Daggula KR, Agarwal R, Shang M, Assi R, Geirsson A, Vallabhajosyula P. Development and Validation of a Predictive Model to Identify Patients With an Ascending Thoracic Aortic Aneurysm. Journal of the American Heart Association. 2021;10:e022102.

8. Obel LM, Diederichsen AC, Steffensen FH, Frost L, Lambrechtsen J, Busk M, Urbonaviciene G, Egstrup K, Karon M, Rasmussen LM, Gerke O, Bovling AS, Lindholt JS. Population-Based Risk Factors for Ascending, Arch, Descending, and Abdominal Aortic Dilations for 60-74–Year-Old Individuals. Journal of the American College of Cardiology. 2021;78:201–211.

9. Pirruccello JP, Lin H, Khurshid S, Nekoui M, Weng L-C, Ramachandran VS, Isselbacher EM, Benjamin EJ, Lubitz SA, Lindsay ME, Ellinor PT. Development of a Prediction Model for Ascending Aortic Diameter Among Asymptomatic Individuals. JAMA. 2022;328:1935–1944.

10. Pirruccello JP, Chaffin MD, Chou EL, Fleming SJ, Lin H, Nekoui M, Khurshid S, Friedman SF, Bick AG, Arduini A, Weng L-C, Choi SH, Akkad A-D, Batra P, Tucker NR, Hall AW, Roselli C, Benjamin EJ, Vellarikkal SK, Gupta RM, Stegmann CM, Juric D, Stone JR, Vasan RS, Ho JE, Hoffmann U, Lubitz SA, Philippakis AA, Lindsay ME, Ellinor PT. Deep learning enables genetic analysis of the human thoracic aorta. Nat Genet. 2021;

11. Collins GS, Reitsma JB, Altman DG, Moons KGM. Transparent Reporting of a multivariable prediction model for Individual Prognosis Or Diagnosis (TRIPOD): The TRIPOD Statement. Annals of Internal Medicine. 2015;162:55.

12. Collins R. UK Biobank Protocol [Internet]. 2007;Available from: https://www.ukbiobank.ac.uk/media/gnkeyh2q/study-rationale.pdf

13. Denny JC, Rutter J, Goldstein DB, Philippakis A, Smoller J, Jenkins G, Dishman E. The “All of Us” Research Program. New England Journal of Medicine. 2019;381:668–676.

14. Rogers IS, Massaro JM, Truong QA, Mahabadi AA, Kriegel MF, Fox CS, Thanassoulis G, Isselbacher EM, Hoffmann U, O’Donnell CJ. Distribution, determinants, and normal reference values of thoracic and abdominal aortic diameters by computed tomography (from the Framingham Heart Study). Am J Cardiol. 2013;111:1510–1516.

15. Pirruccello JP, Rämö JT, Choi SH, Chaffin MD, Kany S, Nekoui M, Chou EL, Jurgens SJ, Friedman SF, Juric D, Stone JR, Batra P, Ng K, Philippakis AA, Lindsay ME, Ellinor PT. The Genetic Determinants of Aortic Distention. Journal of the American College of Cardiology. 2023;81:1320–1335.

16. Khurshid S, Reeder C, Harrington LX, Singh P, Sarma G, Friedman SF, Di Achille P, Diamant N, Cunningham JW, Turner AC, Lau ES, Haimovich JS, Al-Alusi MA, Wang X, Klarqvist MDR, Ashburner JM, Diedrich C, Ghadessi M, Mielke J, Eilken HM, McElhinney A, Derix A, Atlas SJ, Ellinor PT, Philippakis AA, Anderson CD, Ho JE, Batra P, Lubitz SA. Cohort design and natural language processing to reduce bias in electronic health records research. NPJ Digit Med. 2022;5:47.

17. Mbatchou J, Barnard L, Backman J, Marcketta A, Kosmicki JA, Ziyatdinov A, Benner C, O’Dushlaine C, Barber M, Boutkov B, Habegger L, Ferreira M, Baras A, Reid J, Abecasis G, Maxwell E, Marchini J. Computationally efficient whole-genome regression for quantitative and binary traits. Nat Genet. 2021;53:1097– 1103.

18. Sudlow C, Gallacher J, Allen N, Beral V, Burton P, Danesh J, Downey P, Elliott P, Green J, Landray M, Liu B, Matthews P, Ong G, Pell J, Silman A, Young A, Sprosen T, Peakman T, Collins R. UK biobank: an open access resource for identifying the causes of a wide range of complex diseases of middle and old age. PLoS medicine. 2015;12:e1001779.

19. Bycroft C, Freeman C, Petkova D, Band G, Elliott LT, Sharp K, Motyer A, Vukcevic D, Delaneau O, O’Connell J, Cortes A, Welsh S, Young A, Effingham M, McVean G, Leslie S, Allen N, Donnelly P, Marchini J. The UK Biobank resource with deep phenotyping and genomic data. Nature. 2018;562:203.

20. Ge T, Chen C-Y, Ni Y, Feng Y-CA, Smoller JW. Polygenic prediction via Bayesian regression and continuous shrinkage priors. Nature Communications. 2019;10:1776.

21. Karlson EW, Boutin NT, Hoffnagle AG, Allen NL. Building the Partners HealthCare Biobank at Partners Personalized Medicine: Informed Consent, Return of Research Results, Recruitment Lessons and Operational Considerations. J Pers Med [Internet]. 2016 [cited 2020 Jul 14];6. Available from: https://www.ncbi.nlm.nih.gov/pmc/articles/PMC4810381/

22. Kowalski MH, Qian H, Hou Z, Rosen JD, Tapia AL, Shan Y, Jain D, Argos M, Arnett DK, Avery C, Barnes KC, Becker LC, Bien SA, Bis JC, Blangero J, Boerwinkle E, Bowden DW, Buyske S, Cai J, Cho MH, Choi SH, Choquet H, Cupples LA, Cushman M, Daya M, Vries PS de, Ellinor PT, Faraday N, Fornage M, Gabriel S, Ganesh S, Graff M, Gupta N, He J, Heckbert SR, Hidalgo B, Hodonsky C, Irvin MR, Johnson AD, Jorgenson E, Kaplan R, Kardia SL, Kelly TN, Kooperberg C, Lasky-Su JA, Loos RJF, Lubitz SA, Mathias RA, McHugh CP, Montgomery C, Moon J-Y, Morrison AC, Palmer ND, Pankratz N, Papanicolaou GJ, Peralta JM, Peyser PA, Rich SS, Rotter JI, Silverman EK, Smith JA, Smith NL, Taylor KD, Thornton TA, Tiwari HK, Tracy RP, Wang T, Weiss ST, Weng LC, Wiggins KL, Wilson JG, Yanek LR, Zöllner S, North KN, Auer PL, Consortium NT- O for PM (TOPMed), Hematology &amp Topm, Group HW, Raffield LM, Reiner AP, Li Y. Use of &gt;100,000 NHLBI Trans-Omics for Precision Medicine (TOPMed) Consortium whole genome sequences improves imputation quality and detection of rare variant associations in admixed African and Hispanic/Latino populations. bioRxiv. 2019;683201.

23. Altshuler DM, Gibbs RA, Peltonen L, Altshuler DM, Gibbs RA, Peltonen L, Dermitzakis E, Schaffner SF, Yu F, Peltonen L, Dermitzakis E, Bonnen PE, Altshuler DM, Gibbs RA, de Bakker PIW, Deloukas P, Gabriel SB, Gwilliam R, Hunt S, Inouye M, Jia X, Palotie A, Parkin M, Whittaker P, Yu F, Chang K, Hawes A, Lewis LR, Ren Y, Wheeler D, Gibbs RA, Marie Muzny D, Barnes C, Darvishi K, Hurles M, Korn JM, Kristiansson K, Lee C, McCarroll SA, Nemesh J, Dermitzakis E, Keinan A, Montgomery SB, Pollack S, Price AL, Soranzo N, Bonnen PE, Gibbs RA, Gonzaga-Jauregui C, Keinan A, Price AL, Yu F, Anttila V, Brodeur W, Daly MJ, Leslie S, McVean G, Moutsianas L, Nguyen H, Schaffner SF, Zhang Q, Ghori MJR, McGinnis R, McLaren W, Pollack S, Price AL, Schaffner SF, Takeuchi F, Grossman SR, Shlyakhter I, Hostetter EB, Sabeti PC, Adebamowo CA, Foster MW, Gordon DR, Licinio J, Cristina Manca M, Marshall PA, Matsuda I, Ngare D, Ota Wang V, Reddy D, Rotimi CN, Royal CD, Sharp RR, Zeng C, Brooks LD, McEwen JE, The International HapMap 3 Consortium, Principal investigators, Project coordination leaders, Manuscript writing group, Genotyping and QC, ENCODE 3 sequencing and SNP discovery, Copy number variation typing and analysis, Population analysis, Low frequency variation analysis, Linkage disequilibrium and haplotype sharing analysis, et al. Integrating common and rare genetic variation in diverse human populations. Nature. 2010;467:52–58.

24. Li Y, Willer CJ, Ding J, Scheet P, Abecasis GR. MaCH: Using Sequence and Genotype Data to Estimate Haplotypes and Unobserved Genotypes. Genet Epidemiol. 2010;34:816–834.

25. Venner E, Muzny D, Smith JD, Walker K, Neben CL, Lockwood CM, Empey PE, Metcalf GA, Kachulis C, All of Us Research Program Regulatory Working Group, Mian S, Musick A, Rehm HL, Harrison S, Gabriel S, Gibbs RA, Nickerson D, Zhou AY, Doheny K, Ozenberger B, Topper SE, Lennon NJ. Whole-genome sequencing as an investigational device for return of hereditary disease risk and pharmacogenomic results as part of the All of Us Research Program. Genome Med. 2022;14:34.

26. Lim M, Hastie T. Learning Interactions via Hierarchical Group-Lasso Regularization. Journal of Computational and Graphical Statistics. 2015;24:627– 654.

27. DeLong ER, DeLong DM, Clarke-Pearson DL. Comparing the areas under two or more correlated receiver operating characteristic curves: a nonparametric approach. Biometrics. 1988;44:837–845.

28. Pencina MJ, Steyerberg EW, D’Agostino RB. Extensions of net reclassification improvement calculations to measure usefulness of new biomarkers. Stat Med. 2011;30:11–21.

29. Yengo L, Vedantam S, Marouli E, Sidorenko J, Bartell E, Sakaue S, Graff M, Eliasen AU, Jiang Y, Raghavan S, Miao J, Arias JD, Mukamel RE, Spracklen CN, Yin X, Chen S-H, Ferreira T, Ji Y, Karedera T, Lull K, Lin K, Malden DE, Medina- Gomez C, Machado M, Moore A, Rueger S, Group GC-HW, Team 23andMe Research, Program VMV, Initiative) D (DiscovEHR and MCH, Network) eMERGE (Electronic MR and G, Study LC, Center RG, Consortium TP, Group USS, Chasman DI, Cho YS, Heid IM, McCarthy MI, Ng MCY, O’Donnell CJ, Rivadeneira F, Thorsteinsdottir U, Sun YV, Thai ES, Boehnke M, Deloukas P, Justice AE, Lindgren CM, Loos RJF, Mohlke KL, North KE, Stefansson K, Walters RG, Winkler TW, Young KL, Loh P-R, Yang J, Esko T, Assimes TL, Auton A, Abecasis GR, Willer CJ, Locke AE, Berndt SI, Lettre G, Frayling TM, Okada Y, Wood AR, Visscher PM, Hirschhorn JN. A Saturated Map of Common Genetic Variants Associated with Human Height from 5.4 Million Individuals of Diverse Ancestries [Internet]. 2022 [cited 2022 Jan 11]. Available from: https://www.biorxiv.org/content/10.1101/2022.01.07.475305v1

30. Gurdasani D, Carstensen T, Fatumo S, Chen G, Franklin CS, Prado-Martinez J, Bouman H, Abascal F, Haber M, Tachmazidou I, Mathieson I, Ekoru K, DeGorter MK, Nsubuga RN, Finan C, Wheeler E, Chen L, Cooper DN, Schiffels S, Chen Y, Ritchie GRS, Pollard MO, Fortune MD, Mentzer AJ, Garrison E, Bergström A, Hatzikotoulas K, Adeyemo A, Doumatey A, Elding H, Wain LV, Ehret G, Auer PL, Kooperberg CL, Reiner AP, Franceschini N, Maher D, Montgomery SB, Kadie C, Widmer C, Xue Y, Seeley J, Asiki G, Kamali A, Young EH, Pomilla C, Soranzo N, Zeggini E, Pirie F, Morris AP, Heckerman D, Tyler-Smith C, Motala AA, Rotimi C, Kaleebu P, Barroso I, Sandhu MS. Uganda Genome Resource Enables Insights into Population History and Genomic Discovery in Africa. Cell. 2019;179:984–1002.e36.

